# Investigation of antibodies against Chikungunya, Dengue and Zika virus in serum samples from febrile patients and its co-occurrence with malaria in six districts highly endemic for malaria in Mozambique between 2017 – 2018

**DOI:** 10.1101/2022.06.10.22276235

**Authors:** Sádia Ali, Osvaldo Inlamea, Argentina Muianga, Plácida Maholela, John Oludele, Bibiana Melchior, Virgilio Antonio, Vanessa Monteiro, Telma Isaias, Andarusse Sumail, Inocêncio Chongo, Pascoal Alho, Pirolita Mambo, Norbert Heinrich, Eduardo Gudo

## Abstract

**Introduction:** Arboviruses and malaria are both mosquito-borne diseases, with overlapping symptoms and geographic distribution in tropical settings. However, little information is available on the co-occurrence of arboviruses and malaria in areas highly endemic for malaria in Africa. This study was conducted with the aim to determine the frequency of recent Dengue (DENV), Chikungunya (CHIKV) and Zika (ZIKV) infections and their co-occurrence with malaria in six highly endemic districts for malaria in Mozambique.

**Methods:** Blood samples were collected from febrile outpatients between January 2017 and December 2018 and were then tested for Malaria using RDT. Serum samples from these patients were also tested using commercially available ELISA for the presence of IgM antibodies against CHIKV, ZIKV and DENV, as well as NS1 antigen for DENV. Concurrently, a questionnaire was administered to collect socio-demographic characteristics of patients.

**Results:** Of the 906 participants enrolled, IgM antibodies against CHIKV, DENV and ZIKV were identified in 134 (14.8%), 64 (7.4%) and 83 (9.2%) individuals, respectively. Malaria was diagnosed in 56 (6.2%) participants, of which 16 (28.6%) were also positive for IgM anti CHIKV, 1 (1.8%) for DENV-NS1, 3 (5.4%) for IgM anti-DENV and 10 (17.8%) for IgM anti-ZIKV. There was a trend towards an increase in the frequency of IgM anti CHIKV positive samples, from 7.0% in 2014 to 14.8% in 2018 and IgM anti-ZIKV positive samples increased from 4.9% in 2015 to 9.2% in 2018.

**Conclusion:** This study showed an increased frequency of arbovirus in Mozambique thus far, as well as a frequent occurrence of arbovirus among malaria positive patients. This highlighted the urgency for the establishment of sentinel surveillance sites for arboviruses and the need for an integrated management of febrile illnesses in places where arboviruses and malaria are both prevalent.

**AUTHOR SUMMARY:** Arbovirus and malaria share clinical features, which might make the differential diagnosis of acute febrile illnesses significantly difficult, leading to frequent over diagnosis of malaria and under diagnosis of arbovirus in places where both co-occur. In Mozambique and other sub-Saharan countries, epidemiological data on the burden and temporal trend of arbovirus, as well as on co-occurrence with malaria are Chikungunya virus (CHIKV) and Dengue virus (DENV) and its co-occurrence with malaria among 609 febrile patients in six health centres located in five provinces in Mozambique. Recent CHIKV, DENV and ZIKV infection, as measured by presence of IgM antibodies, were found in 14.8%, 7.4% and 9.2% of participants and among 28.6%, 5.4% and 17.8%, of malaria positive patients. These findings suggest that arboviruses are frequent among malaria and non-malaria febrile patients, reinforcing the need for increased awareness of arbovirus in the management of acute febrile illness.

## INTRODUCTION

Chikungunya (CHIKV), Dengue (DENV) and Zika virus (ZIKV) are arboviruses transmitted mostly by *Aedes aegypti* and *Aedes albopictus* (1), the first having shown to be abundant in many geographical places in southern Africa, including Mozambique (2-4). Many of these African countries are also highly endemic for malaria, which might make the differential diagnosis for acute febrile illness somewhat difficult, leading to frequent underdiagnosis of arboviral infections and overdiagnosis of malaria. Using clinical suspicions as part of the diagnostic of arbovirus is challenging because of the absence of specific symptoms, aggravated by similarity of its symptoms with those of malaria (5-7). Frequent co-infections of arboviruses and malaria among febrile patients have been described where both pathogens co-occur (8). Mozambique is among the six countries accounting for more than half of all malaria cases and deaths worldwide (9)(10). Besides this, a study conducted among acute febrile patients in 22 health facilities in the country demonstrated that almost half of the patients who had received anti-malarial drugs, received a negative malaria result, highlighting poor management of febrile illness (11) and the lack of information regarding the co-occurrence of malaria and arboviruses in Mozambique.

In this context, this study was conducted with the aim of investigating the frequency of recent infections by CHIKV, DENV and ZIKV and their co-occurrence with malaria, among febrile patients in 6 selected health facilities. This study is important to provide data on the co-infection of arboviruses and malaria, which is relevant to guide improvement of case management for febrile illness in Mozambique.

## MATERIALS AND METHODS

### Study design and settings

A cross-sectional study was conducted among outpatients with acute febrile illness attending 6 health facilities (Polana Caniço, Massingir, Eduardo Mondlane, Coalane and Natite Health facilities and Caia Rural Hospital), distributed across 6 different provinces, between January 2017 and December 2018.

Mozambique is situated in the southeast coast of Africa and has a tropical climate with two distinct seasons, the rainy season that begins in October and ends in March and the dry season, from April to September. The relative humidity is high, ranging between 70 and 80% and the average annual precipitation is estimated at 600 mm, varying between 500 and 900 mm. The country is administratively divided into 11 provinces and 164 districts and the total population is estimated at 31 million, of which approximately 70% live in rural areas (12).

### Participant recruitment and procedures

Participants who were older than 5 years, with acute febrile illness (temperature ≥ 38°C, measured or self-reported) and able to provide written informed consent were eligible. A questionnaire was applied to obtain demographic, socioeconomic and clinical data. All participants were screened using a Malaria Rapid Diagnostic Tests (RDT) during their medical appointment at the time of enrolment, using the national algorithms for Malaria diagnostic. Pregnant women and those individuals with psychiatric diseases, as well as patients with a readily identifiable focus of infection were excluded.

### Ethics statement

Ethical approval for this study was obtained from the Mozambican National Bioethics Committee for Health (Ref. No. 487/CNBS/2017). A written informed consent was obtained from all study participants and from parents or guardians of children under the age of 18. All questionnaires were handled anonymously and confidentially.

### Laboratory testing

All participants were initially tested for malaria using a RDT specific for *Plasmodium falciparum*, according to the manufacturer’s instructions (SD BIOLINE Malaria Ag P.f, SD. Inc. Yongin-si, Gyeonggi-do, South Korea). The serum samples were shipped under a cold chain to the Virology Reference Laboratory at INS in Maputo Province, where they were tested in duplicate for human immunoglobulin M (IgM) antibodies against CHIKV and ZIKV using a commercially available enzyme-linked immunosorbent assay (ELISA) (Euroimmun Lübeck, Germany). Regarding DENV, samples were also screened for the non-structural protein 1 (NS1) antigen and IgM antibodies against DENV also using commercially available ELISA kits (Panbio™ Abbott, Australia). The interpretation of the test results was based on the manufacturer instructions.

### Statistical analysis

Socio-demographic and serological data were entered first into an electronic database developed using EPI Info™ CDC, followed by a double entry which was done by a different individual. Data were analysed using the R package and for descriptive statistics for all continuous variables the confidence interval was calculated (95%) as well as the interquartile ranges (IQR). Comparison between proportions of categorical variables was done using The Chi-Square test. All tests were considered statistically significant at p < 0.05.

## RESULTS

### Demographic characteristics of participants

From January 2017 to December 2018, a total of 906 febrile patients were recruited from the following six health facilities (HF): Polana Caniço General Hospital in Maputo City (n = 137), Massingir Health Center in Gaza Province (n = 117), Caia District Hospital in Sofala province (n = 120), Eduardo Mondlane Health Center in Chimoio Province (n = 246), Coalane Health Center in Zambezia Province (n = 211) and Natite Health Center in Cabo Delgado Province (n = 75). Of those, 560 (62.5%) were female (Figure 1) and the median age of participants was 25 years old, most of them aged between 18 and 24 years (258/906; 28.8%) (Table 1).

**Figure 1:**
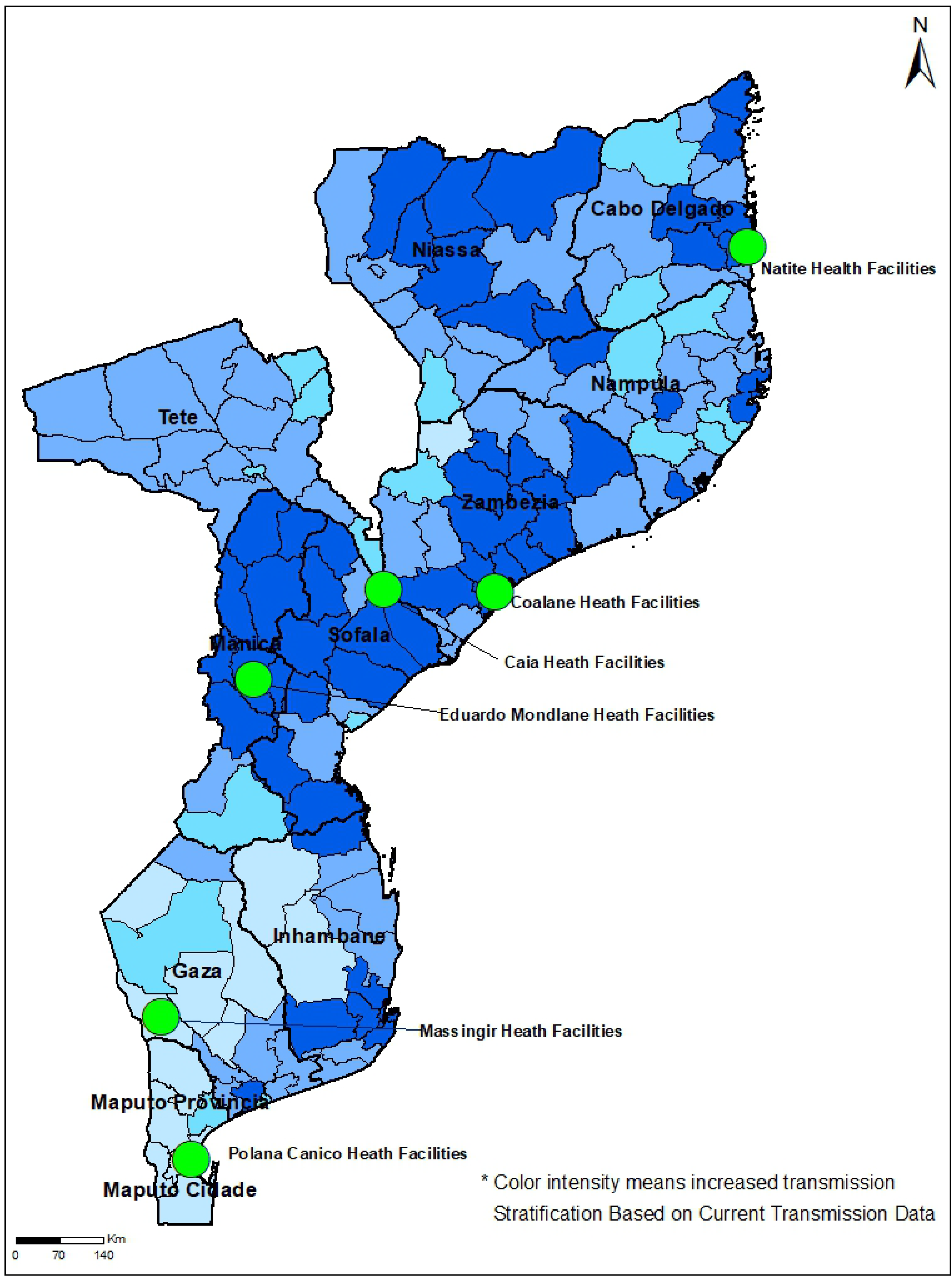
Malaria burdened districts and the geographical localization of the 6 study sites. The figure depicts the of malaria burdened district (in blue colour gradient) and the geographical localization of the 6 study sites (green circles)

**Table 1:**
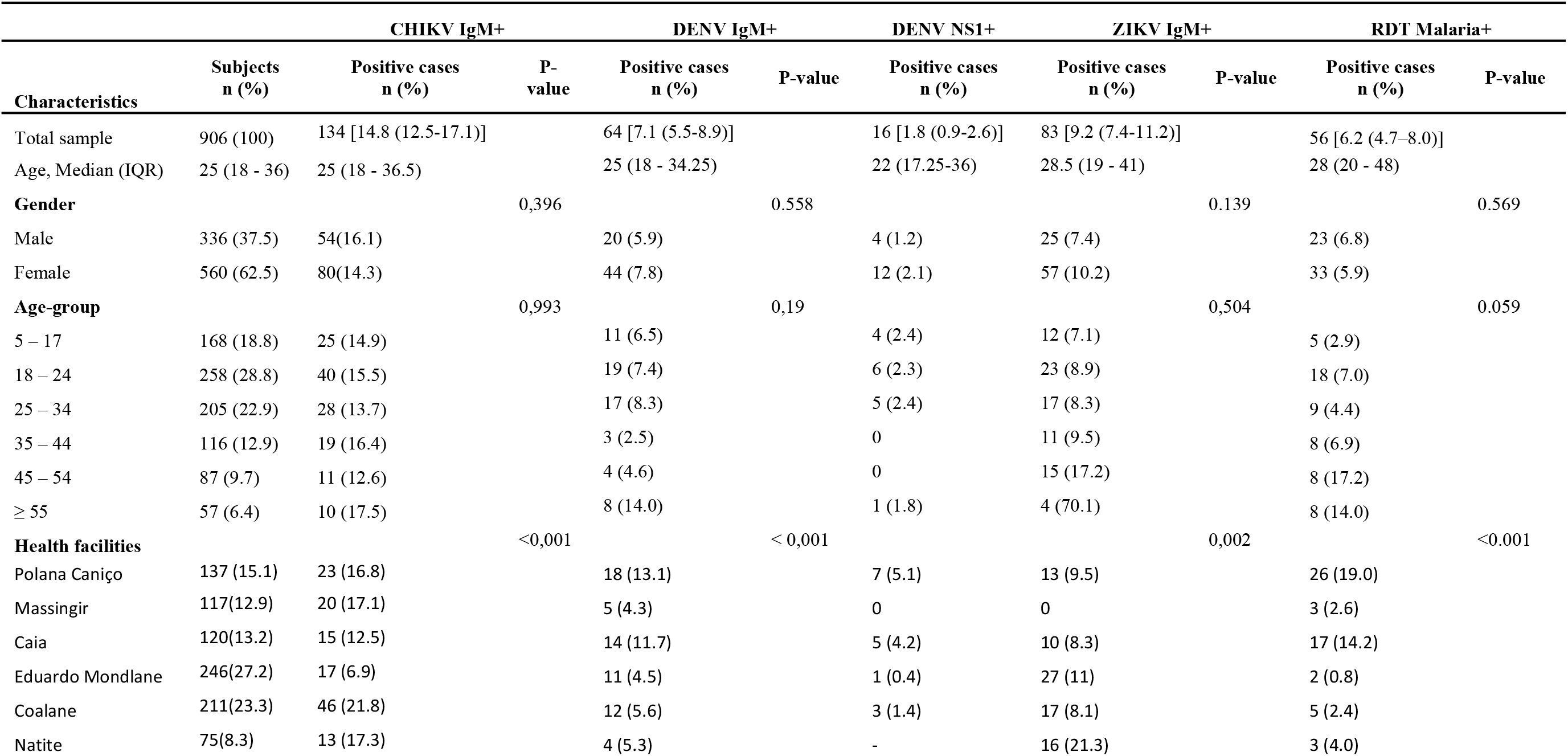
Demographic characteristics of the participants.

### IgM anti-CHIKV antibodies

A total of 134 out of 906 (14.8%; 95%CI: 12.5-17.1) patients tested positive for IgM anti-CHIKV antibodies with a median age of 25 (IQR: 18.0 – 36.5) years, although the highest frequency was reported in participants aged more than 55-year-old (10/57; 17.5%). No statistically significant difference was found in the frequency of IgM anti-CHIKV antibodies in female which was higher when compared to male (p = 0.396). In terms of geographical distribution, there were statistically significant differences in the frequencies of IgM anti-CHIKV antibodies between HF (p < 0.000), Coalane being the HF with the highest frequency (Table 1).

### IgM anti-DENV antibodies and DENV NS1

Of the 906 participants enrolled, 64 (7.1%; 95%CI: 5.5-8.9) tested positive for IgM anti-DENV antibodies, with a median age of 25 (IQR: 18.0 – 34.3) years. No statistically significant difference was found in the frequency of IgM anti-DENV positive cases between male and female (p = 0.558), and between age categories (p = 0.191). There were, however, differences in the frequency of IgM anti-DENV antibodies between HFs, the highest being found in Polana Caniço General Hospital (18/137; 13.1%), followed by Caia District Hospital (14/120; 11.7%) (p < 0.001). Frequency of DENV NS1 among participants was 16/906 (1.8%; 95% CI: 0.9 – 2.6) with a median age of 22 (IQR:17.3 – 36.0) years (Table 1).

### IgM anti-ZIKV antibodies

A total of 83 out of 906 participants (9.2%; 95%CI: 7.4-11.2) were positive for IgM anti-ZIKV, with a median age of 28.5 (IQR: 19 – 41) years. No statistically significant difference was found between male and female (57/606; 10.2% for male versus 25/336; 7.4% for female, p = 0.139), although there was a statistically significant difference in the frequency IgM anti-ZIKV antibodies between HFs (p = 0.002), the highest being observed in Natite Health Center (16/75; 21.3%) followed by Eduardo Mondlane Health Center (27/246; 11.0%) (see Table 1).

### Malaria

Among the enrolled patients, 56/906 (6.2%; 95%CI: 4.7-8.0) were malaria RDT positive, with a median age of 28 (IQR: 20 – 48) years. No statistically significant difference was found between male and female (23/336; 6.8% for male versus 33/560; 5.9% for female, p = 0.569) (Table1). Proportion of malaria positive cases was significantly different among HFs (p < 0.001) and the highest frequency was reported at Polana Caniço General Hospital (26/137; 19.0%).

### Monthly distribution of CHIKV, DENV, ZIKV and malaria cases

An assessment of the monthly trend of positivity for IgM anti-CHIKV, DENV and ZIKV antibodies was performed over a 24-month period. Figure 2 shows a progressive increase in the number of febrile patients and the proportion of IgM anti CHIKV, DENV and ZIKV antibodies as well as of malaria positive cases during the 2 years of this study (Figure 2).

**Figure 2:**
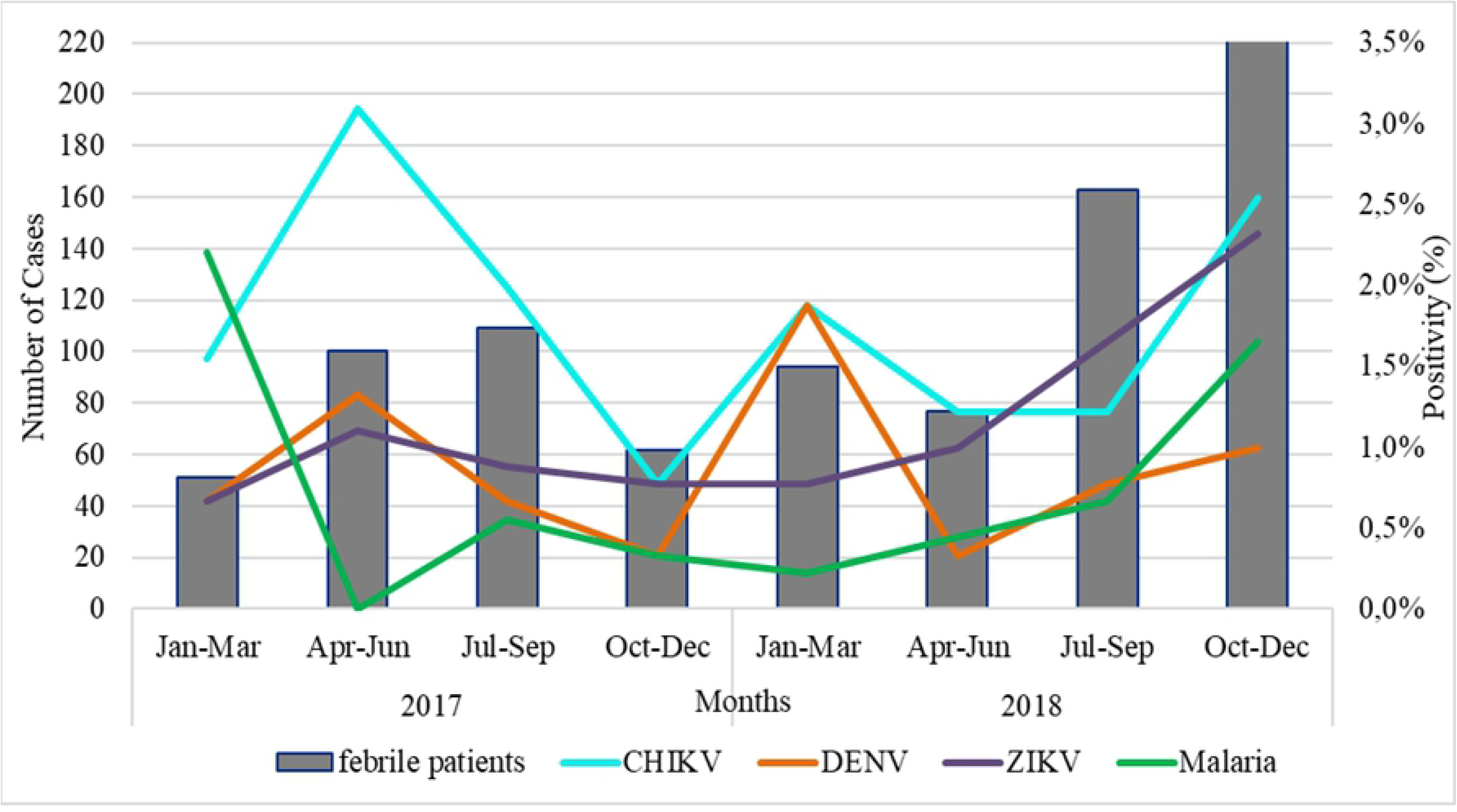
Frequency (%) of cases of CHIKV, DENV, ZIKV recent infections and number (n) of malaria infections among acute febrile individuals between January 2017 and December 2018.

### Temporal trend of arboviruses in Mozambique

Table 2 shows a comparison of the frequency of IgM antibodies against arbovirus over the years using data from the current and from previous studies. There has been an increase in the frequency and distribution of IgM antibodies against CHIKV from 6.0% in 2008 to 14.8% in the current study and increase in the frequency of IgM antibodies against ZIKV from 4.9% in 2019 to 9.2% in the current study. Frequency of IgM antibodies against DENV was quite heterogeneous throughout the years, with no specific trend.

**Table 2:** Comparison of frequency of IgM antibodies against CHIKV, DENV and ZIKV in different studies between 2009 and 2018

| Study characteristic | Sample size | Sample Source | Timeframe (duration) | Source population | Country Regions | IgM anti-CHIKV | IgM anti-DENV | IgM anti-ZIKV |
| --- | --- | --- | --- | --- | --- | --- | --- | --- |
| Current study | 906 | Sentinel surveillance | Jan 2017 to Dec 2018 | 6 health facilities - | North, Central and South Regions | 134/906 (14.8%) | 64/906 (7.4%) | 83/906 (9.2%) |
| *Mugabe et al [10]* | 163 | Serological investigation | Feb to Jun 2016 | 5 health units in Quelimane, Zambezia | Central Region | 17/163 (10.4%) | 1/104 (0.9%) | - |
| *Oludele et al [13]* | 192 | Post-outbreak Surveillance | Jan 2015 to Mar 2016 | Pemba and Nampula | North Region | - | 39/192 (20%) | - |
| *Muianga et al [11]* | 114 | Outbreak investigation | Mar to April 2014 | Pemba, Cabo Delgado | North Region | 8/114 (7.0%) | 46/120 (38.3%) | - |
| *Antonio et al* [12] | 895 | Serological testing of serum samples from the INS biobank | 2009 to 2015 | 127 Health facilities | North, Central and South Regions | 54/895 (6.0%) | - | - |
| *Chelene et al* [14] | 850 | Serological testing of serum samples from the INS biobank | 2009 to 2015 | 127 health facilities | North, Central and South Regions | - | - | 42/85 (4.9%) |

### Co-occurrence of IgM antibodies against CHIKV, DENV, ZIKV and malaria

The frequencies of co-occurrence of IgM anti-CHIKV, DENV and ZIKV antibodies among malaria positive participants are described in Table 3 which shows that the higher frequency of co-infection was 16 (28.6.9%; 95%CI: 17.3 – 42.2) for IgM anti CHIKV, followed by 10 (17.8%; 95%CI:8.9 – 30.4) for IgM anti-ZIKV, 3 (5.4%; 95%CI:1.1 – 14.9) for IgM anti-DENV and 1 (1.8%; 95%CI:0.45 – 9.5) for DENV-NS1.

**Table 3:** Distribution of co-occurrence of IgM antibodies against CHIKV, DENV and ZIKV with malaria.

| Combination of biomarkers | Positive cases<br>(malaria positive = 56) | % (95% CI) |
| --- | --- | --- |
| IgM anti-CHIKV + Malaria | 16 | 28.6 (17.3 – 42.2) |
| IgM anti-DENV + Malaria | 3 | 5.4 (1.1 – 14.9) |
| DENV NS1 + Malaria | 1 | 1.8 (0.45 – 9.5) |
| IgM anti-ZIKV + Malaria | 10 | 17.9 (8.9 – 30.4) |

## DISCUSSION

This study represents the largest and the most recent cross-sectional serological investigation of CHIKV, ZIKV and DENV in serum from febrile patients in Mozambique. This also represents the largest study to address co-occurrence of recent arbovirus infections with malaria. We firstly described the sero-epidemiology of arbovirus and found a frequency of anti-CHIKV IgM antibodies of 14.8%, which is the highest reported in Mozambique thus far. Previous studies found lower frequencies of IgM anti-CHIKV antibodies (13, 14), suggesting a recent increase in the frequency of CHIKV in the country. The global distribution of CHIKV shows the increase in frequency and an alarming rate of expansion to new areas (15). The reasons that drove an increase in the frequency of CHIKV are yet to be determined, but can be attributed to higher abundance of its mosquito vector, as well as changes in the human behavior and environmental and climate changes that create a better environment for the growth of *Aedes* mosquitoes (16, 17). In this study, the higher positivity of IgM antibodies against CHIKV was found in Coalane Health Center, situated in central Mozambique, which has been corroborated by previous studies conducted in the country (13, 18).

Regarding DENV, this study found a frequency of IgM anti-DENV antibodies of 7.1%, which is slightly lower than that reported in a previous outbreak of DENV in Cabo Delgado province in northern Mozambique (19). This suggests a well-established endemic transmission of DENV in Mozambique, with periodic epidemic transmission that facilitate the emergence of outbreaks as shown in other countries in the continent (16, 17).

Data on ZIKV epidemiology are scarce in the country. This study reported a frequency of IgM anti-ZIKV of 9.2%, which is higher than the prevalence of 4.9% found in a recent retrospective study conducted among febrile children aged 1 to 15 years old in Mozambique (20), suggesting that the frequency of ZIKV is increasing in the country. Similar to CHIKV, we believe that several factors are behind the potential increase in the frequency of ZIKV, such as human behaviors and climate and environmental changes (21-23) (24). The few data available in the continent on ZIKV suggests that the virus is endemic in Africa.

In general, there is a growing consensus that climate change is driving a rapid increase in the burden of arbovirus and other mosquito borne diseases (23), as it affects vector distribution and abundance, life cycle, survival, biting, pathogen incubation rates, and vector competence. (17, 23, 25-27). Besides this, *Aedes aegypti* is well distributed in Mozambique (2, 3, 28).

On the other hand, the frequency of extreme climate events such as cyclones and floods have been increasing significantly, which also increases human mobility and causes displacement of a large number of population and communities (29, 30). These massive population movements contribute to changes in the epidemiology of infectious diseases, including arboviruses (26, 27).

The increased burden of arbovirus in Mozambique and the overlap of clinical presentation with malaria add complexity on overall fever management protocols, despite the few studies addressing co-occurrence or co-infection of malaria and arboviruses. In this study, we investigated the co-occurrence of biomarkers of recent arbovirus infections, measured by the presence of IgM antibodies against arboviruses among malaria positive patients.

Data from this study found a high frequency of IgM antibodies against CHIKV (28.6%) and ZIKV (17.8 %), while frequency of IgM antibodies against DENV (4.9%), as well as frequency of DENV-NS1 (1.8%) were lower, among malaria positive patients. The higher frequency of IgM antibodies against CHIKV and ZIKV among malaria positive cases as compared to that found among febrile patients is a matter for further investigation but might suggest an overlap in the circulation of malaria, CHIKV and ZIKV in Mozambique. This study found a higher frequency of co-occurrence of IgM anti-CHIKV and malaria as compared to previous studies conducted in the country (13, 20), as well as in other countries in the continent (31-33).

Regarding co-occurrence of recent ZIKV infection and malaria, this study found a frequency of co-occurrence of 17.9% (10/56), as similarly reported in other countries in Africa (32, 34).

Regarding co-occurrence between recent DEV infection and malaria, the frequency of occurrence of IgM anti-DENV among malaria positive patients found in this study (4.9%) was similar to that reported in Nigeria (35) and Tanzania (33).

Frequency of co-occurrence of malaria and biomarkers of recent CHIKV, ZIKV or DENV infection found in this study suggest that arbovirus is silently circulating among malaria and non-malaria febrile patients, which reinforces the need for increased awareness of arbovirus among clinicians. Lack of awareness leads to under diagnosis of arboviruses and over diagnosis of malaria, which consequently causes an over prescription of anti-malarian treatment and also a mismatch of public health policy. This also highlights the urgent need to incorporate arboviruses as part of the differential diagnosis of acute fever in Mozambique (36). Laboratory testing for arbovirus is mostly unavailable in Africa, which contributes significantly to under diagnosis of arbovirus and over diagnosis of malaria.

Data from this study showed that half (53.5%) of the malaria cases were seropositive for IgM antibodies against either of DENV, CHIKV and ZIKV, highlighting the high frequency of co-occurrence and co-infection of malaria with arbovirus infections, which has been shown to be heterogeneous in Africa (31, 37). A study conducted in Nigeria found a frequency of co-occurrence of malaria and recent arbovirus infection of 26.3% (10/38) and a frequency of 1.8% (11/600) among febrile patients has been reported in Ghana (35, 38).

The clinical implication of co-occurrence of arbovirus and malaria is still unknown and further studies should be conducted in the future given its increased frequency.

We would like to acknowledge some of the limitations of our study. First, this study was based on serologic tests using commercial ELISA reagents and there is potential cross-reaction of CHIKV with other Alphaviruses, such as O’nyong nyong virus, while ZIKV and DENV are both Flavivirus and can cross react among each other. Regarding, CHIKV ELISA, a previous study conducted in Quelimane city found a 100% agreement between this ELISA and neutralization assay (18), showing that this ELISA essay is highly specific. On the other hand, the use of IgM antibodies against arboviruses allows comparability as most of the previous studies conducted in the country and continent detected IgM antibodies as a proxy of recent infection. A second limitation of this study is the fact that it was conducted in sentinel sites across the country and not in a countrywide context, which limits generalization of the results.

## CONCLUSION

This study represents the largest and most recent cross-sectional descriptive serological investigation of arboviruses among febrile patients in Mozambique and its co-occurrence with malaria. Data from this study show an increased frequency of arboviruses in Mozambique which highlights the need for urgent establishment of sentinel surveillance for these pathogens. The study also found a frequent co-occurrence of malaria and recent infections of CHIKV, DENV and ZIKV among febrile patients, which emphasizes the need of increased awareness on arboviral infection in the differential diagnosis of non-malaria febrile patients.

## Data Availability

All relevant data are within the manuscript and its Supporting Information files.

## Supporting information

**S1 Appendix: Tendency of malaria cases, 2017 to 2018**. a) cases from Natite HF, Pemba city; b) cases from Coalane HF, Quelimane; c) cases from Eduardo Mondlane HF, Chimoio City; d) cases from Caia Hospital, Caia district; e) cases from Massingir HF, Massingir district; and f) cases from Polana Canico Hospital, Maputo city

**S2 Appendix: Monthly distribution of aggregated data of malaria report on the study sites and the data from our study participants**

### ACKNOWLEDGEMENTS

The authors are grateful to the management of all 6-health facilities where the samples were collected and used in this study. We thank all staff from Virology Laboratory at the National Institute of Health for their collaboration during sample testing.

## AUTHOR CONTRIBUTION

**Conceptualization:** Sádia Ali, Argentina Muianga, Vanessa Monteiro, Osvaldo Inlamea, Eduardo Samo Gudo, Norbert Heinrich

**Formal analysis:** Sadia Ali, Pirolita Mambo, Osvaldo Inlamea, Norbert Heinrich, Eduardo S. Gudo

**Funding acquisition:** Osvaldo Inlamea, Eduardo Samo Gudo,

**Investigation and Methodology:** Argentina Muianga, Telma Isaias, John Oludele J, Plácida Maholela, Andarusse Sumail, Virgilio S. António, Inocêncio Chongo, Bibiana Melchior, Pascoal Alho,

**Project administration;** Sádia Ali, Argentina Muianga, Vanessa Monteiro, Osvaldo Inlamea

**Writing_original draft:** Sádia Ali, Osvaldo Inlamea, Plácida Maholela

**Writing-review & editing:** Eduardo Samo Gudo, Norbert Heinrich

